# Global Stroke Burden from Metabolic Risks Across Demographics: Findings from the 2021 Global Burden of Disease Study

**DOI:** 10.1101/2024.08.06.24311583

**Authors:** Song Xue, Guoqing Wu

## Abstract

**Background:** Stroke is the second leading cause of death and the primary cause of disability worldwide. Metabolic risks are major contributors to stroke. The global trends in metabolic risk-related stroke from 1990 to 2019, and the differences in mortality and DALYs across various demographic factors, remain unclear.

**Methods:** All analyses were based on rates derived from the GBD2021 results (https://vizhub.healthdata.org/gbd-results/). Data were stratified by gender, region, and age. Joinpoint software was used to perform regression analysis of the average annual percent change (AAPC) and its 95% confidence interval to analyze trends from 1990 to 2019. Excel, PowerPoint, and R software were used for plotting and analysis, with p<0.05 considered statistically significant.

**Results:** From 1990 to 2019, the average annual percent change (AAPC) for age-standardized rates (ASR) of DALYs was -1.70% (−1.81%, -1.58%), and for mortality, the AAPC was -1.57% (−1.68%, -1.46%). As the SDI increased, both the ASR of DALYs and mortality in 2019 showed a significant decline. The AAPC from 1990 to 2019 also exhibited a downward trend with increasing SDI levels. The DALYs and mortality rates of metabolic risk-related stroke predominantly affected individuals aged 75 and above, with a lesser impact on those under 55. For both genders, the 10-55 and 50-74 age groups had the highest DALYs and mortality rates due to metabolic-related intracerebral hemorrhage. For those aged 75-84 and over 85, ischemic stroke was the leading subtype of metabolic-related stroke contributing to DALYs and mortality rates.

**Conclusion:** This is the first retrospective study on metabolic risk-related stroke on a global scale, summarizing its temporal trends and demographic distribution characteristics. Effective public health strategies are needed to address these disparities and continue reducing the global burden of metabolic risk-related strokes.

## Introduction

Stroke is the second leading cause of death and the primary cause of disability worldwide. ^1^ Over the past three decades, the global burden of stroke has seemingly decreased, primarily reflecting declines in high-income countries. ^2^ In contrast, the burden of stroke has rapidly increased in low- and middle-income countries, ^3^ with variations in age and gender compositions contributing to differences in stroke burden across countries. ^4^

Metabolic factors are key contributors to stroke risk. ^5^ Elevated fasting blood glucose levels, ^6^ high low-density lipoprotein cholesterol, ^7^ high systolic blood pressure, ^8^ elevated body mass index, ^9^ low bone mineral density, ^10^ and impaired renal function ^11^ significantly increase stroke risk. However, comprehensive epidemiological information on stroke burden related to metabolic risk factors remains limited.

In this study, we extracted epidemiological data on stroke related to metabolic risks from the Global Burden of Disease (GBD) 2021 study. The GBD study is a collaborative research project between the World Health Organization (WHO) and many academic institutions that aims to assess the burden of disease and injury at the global, regional and national levels. ^12^ The data of GBD 2021 study were stratified by location, gender, age, and Socio-Demographic Index (SDI) status, and we further analyzed the trends in disease burden over the demographics and time dimensions. This study represents the first global epidemiological analysis specifically addressing stroke related to metabolic factors, providing valuable insights for developing prevention strategies tailored to the metabolic risk profiles of different countries and regions.

## Methods

### 2.1 Data Sources

The GBD 2021 study provided a systematic and comprehensive assessment of diseases and injuries worldwide from 1990 to 2021. ^13^ Data sources included field surveys, censuses, vital registration data, and other health-related data sources. ^12^ The GBD study followed the Guidelines for Accurate and Transparent Health Estimates Reporting (GATHER). ^14^

The results were available from the GBD online results tool (https://vizhub.healthdata.org/gbd-results/) and could be viewed interactively via the GBD compare tool (https://vizhub.healthdata.org/gbd-compare/).).

### 2.2 Exposure and Outcomes

There were three primary risk factors in the GBD study (Supplemental Table 1). Metabolic risk was a primary risk factor, including high fasting plasma glucose, high LDL cholesterol, high systolic blood pressure, high body mass index, low bone mineral density, and impaired kidney function. (Supplemental Table 1) Stroke was a third-level disease in the GBD study, which included three fourth-level diseases. The definition of each disease could be found in previous research. ^15^

SDI, a composite indicator of social and economic factors influencing health outcomes in each region, was calculated by GBD 2021 for each country. ^16^ SDI was categorized into five levels: low, low-middle, middle, high-middle, and high based on GBD result Website (https://vizhub.healthdata.org/gbd-results/). The 204 countries were classified into different SDI regions based on their SDI scores: low SDI (0– 0.455), low-middle SDI (0.456–0.608), middle SDI (0.609–0.690), high-middle SDI (0.690–0.805), and high SDI (0.806–1). ^17^

### 2.3 Data Analysis

To avoid the potential impact of the COVID-19 pandemic ^18,19^ on the results, we limited our analysis to data from 1990 to 2019. Age-standardized rates (ASR) of DALYs and deaths were used to compare the effect of metabolic-related stroke in different years, sexes, regions, and age groups. ^20^ Unless otherwise specified, DALYs and mortality rates were age-standardized and reported per 100,000 persons.

The calculation of time trend was based on the Average Annual Percent Change (AAPC) indicator, ^21^ AAPCs were calculated using the Joinpoint software (Version 5.2.0). ^22^ For calculating the AAPC from 1990 to 2019, we selected 0 joinpoint index.

To determine the proportion of strokes attributable to various risk factors, we utilized the GBD results to obtain DALYs and mortality rates for each risk factor and calculated the proportion by dividing the value for each specific risk factor by the total. Statistical analysis and graphical mapping were conducted using Excel 2020, PowerPoint 2020, and R software (version 4.1.1), with packages ggplot2, ^23^ dplyr, ^24^ countrycode, ^25^ and scatterpie. ^26^ Confidence intervals were two-tailed at 95%, and p-values < 0.05 were considered statistically significant.

## Results

### 3.1 Global Burden of Stroke Attributable to Metabolic Risks

In 2019, the age-standardized DALYs rate due to metabolic risk-related stroke globally was 1321.94 (1104.76, 1507.9) and the mortality rate was 62.73 (52.26, 71.7), both showing significant declines compared to 1990 (Table 1). From 1990 to 2019, the AAPC for age-standardized DALYs was -1.70% (−1.81%, -1.58%), and for mortality rate, the AAPC was -1.57% (−1.68%, -1.46%) (Table 1). Geographically, the world was divided into 33 regions (Table 1). In 2019, Southeast Asia had the highest age-standardized mortality rate (ASMR) of 116.21 (98.02, 133.41) and the highest age-standardized DALYs rate of 2500.54 (2104.75, 2894.53) (Table 1). Except for Southern Africa and Southern Sub-Saharan Africa, the other 31 regions showed significant declines in both rates from 1990 to 2019, with AAPCs less than 0 (Table 1).

**Table 1:**
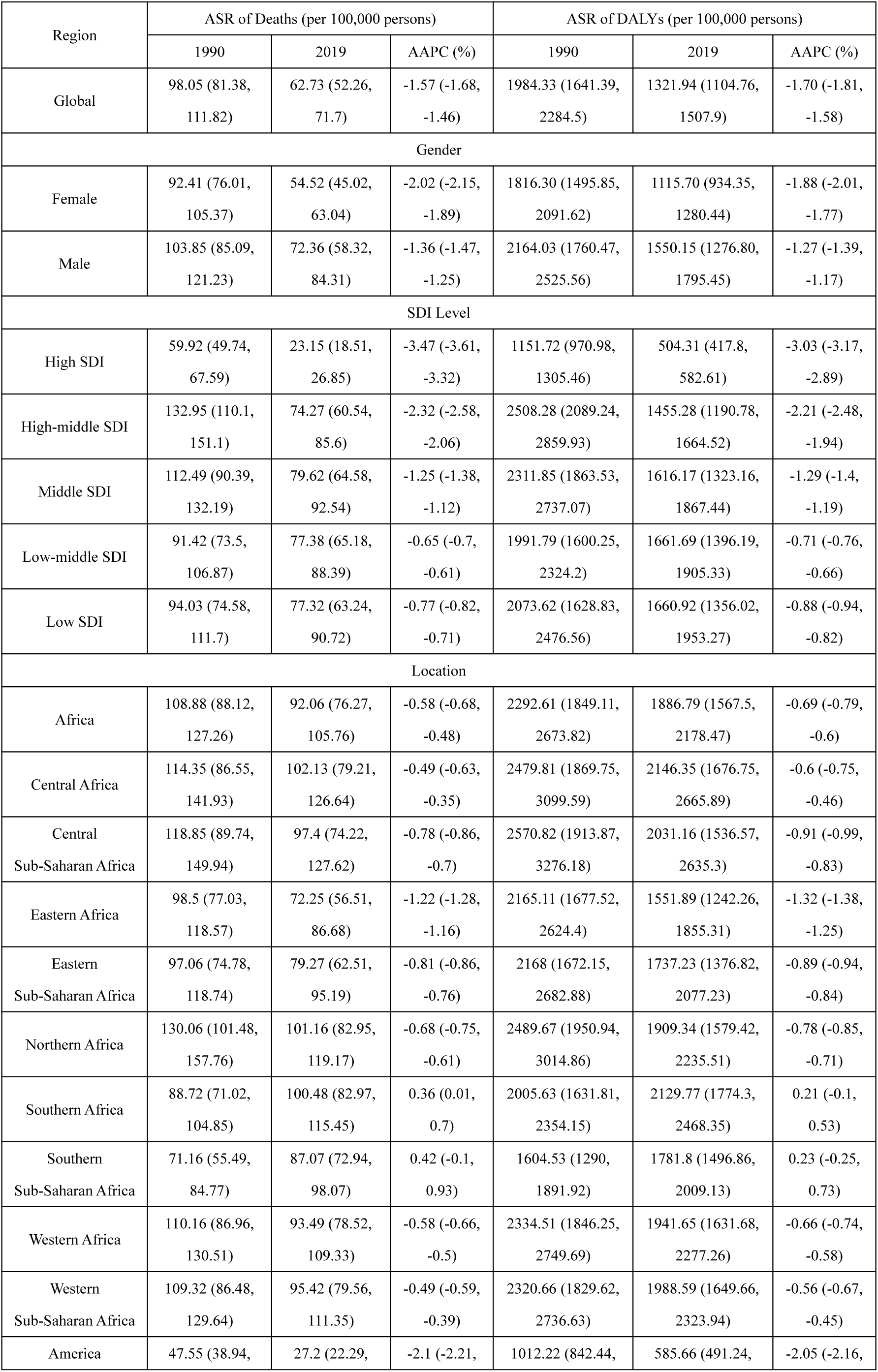
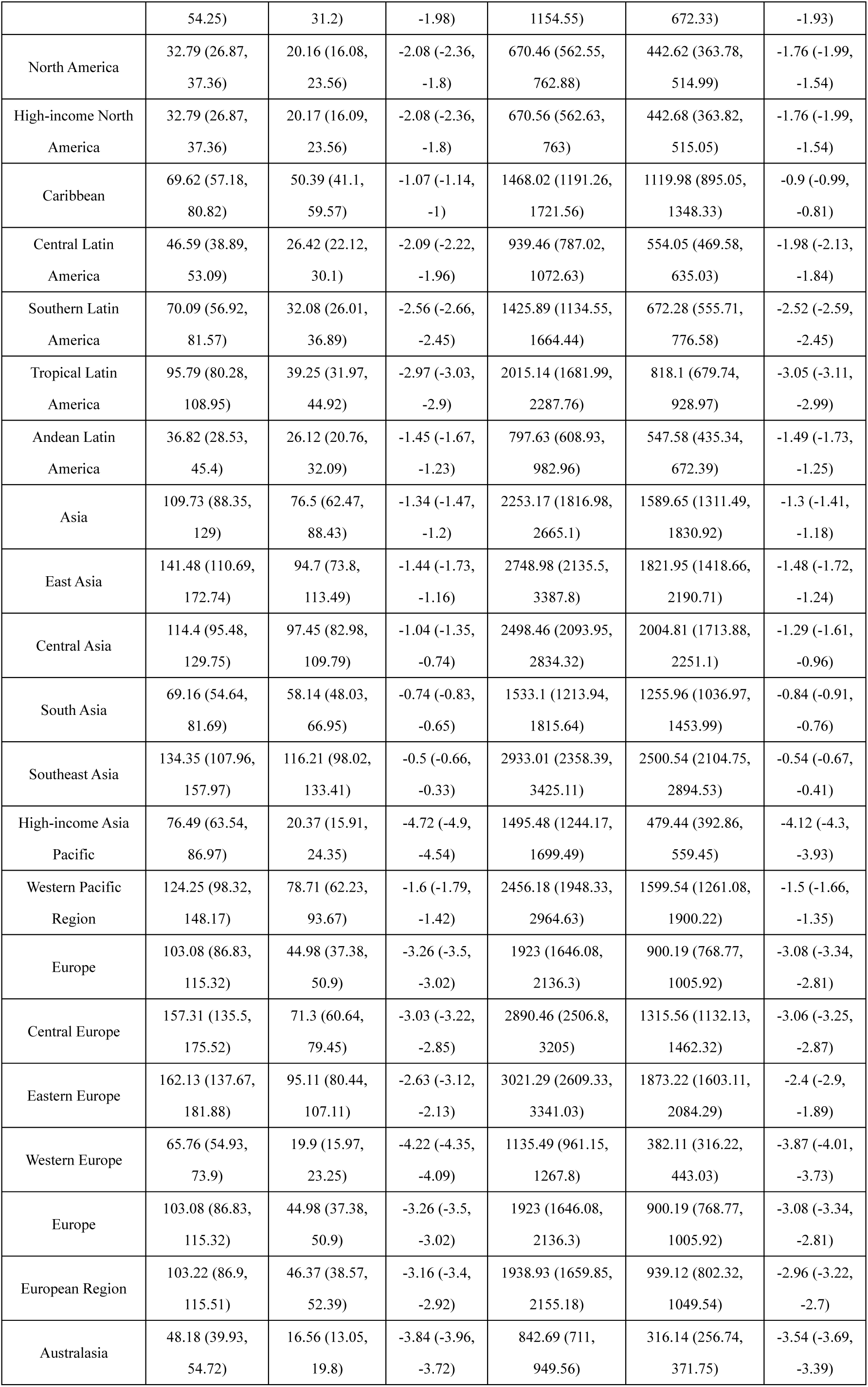
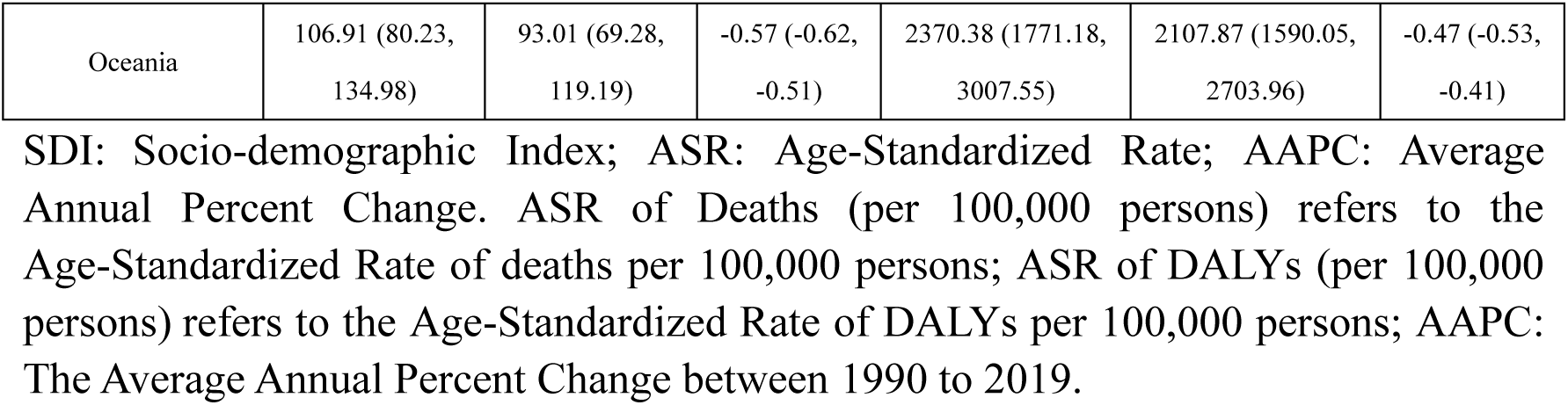
The Global Distribution of Metabolic-related Stroke from 1990 to 2019.

At the country level (Supplemental Table 2, Figure 1), the Republic of Nauru had the highest rates in 1990 with an ASR of DALYs 4759.53 (3678.31, 6011.55) and the Republic of Serbia had an ASMR of 252.39 (212.12, 290.61). In 2019, the highest rates were again in the Republic of Nauru with a DALYs rate of 4527.26 (3412.25, 5837.06) and North Macedonia with a mortality rate of 240.36 (199.66, 277.11). Singapore had the lowest ASMR in 2019 at 9.48 (7.39, 11.45), and the Swiss Confederation had the lowest DALYs ASR at 234.72 (186.91, 277.78) (Supplemental Table 2, Figure 2). A total of 170 countries showed significant declines in DALYs rates, and 165 countries showed significant declines in mortality rates (Supplemental Table 2, Figure 2). The Republic of Korea had the most pronounced decrease in DALYs rate with an AAPC of -6.55% (−6.92%, -6.28%), and the Republic of Estonia had the most pronounced decrease in mortality rate with an AAPC of -6.66% (−7.31%, -6.01%). Conversely, the Kingdom of Lesotho exhibited the most significant increase in both DALYs and mortality rates (Supplemental Table 2, Figure 2).

**Figure 1:**
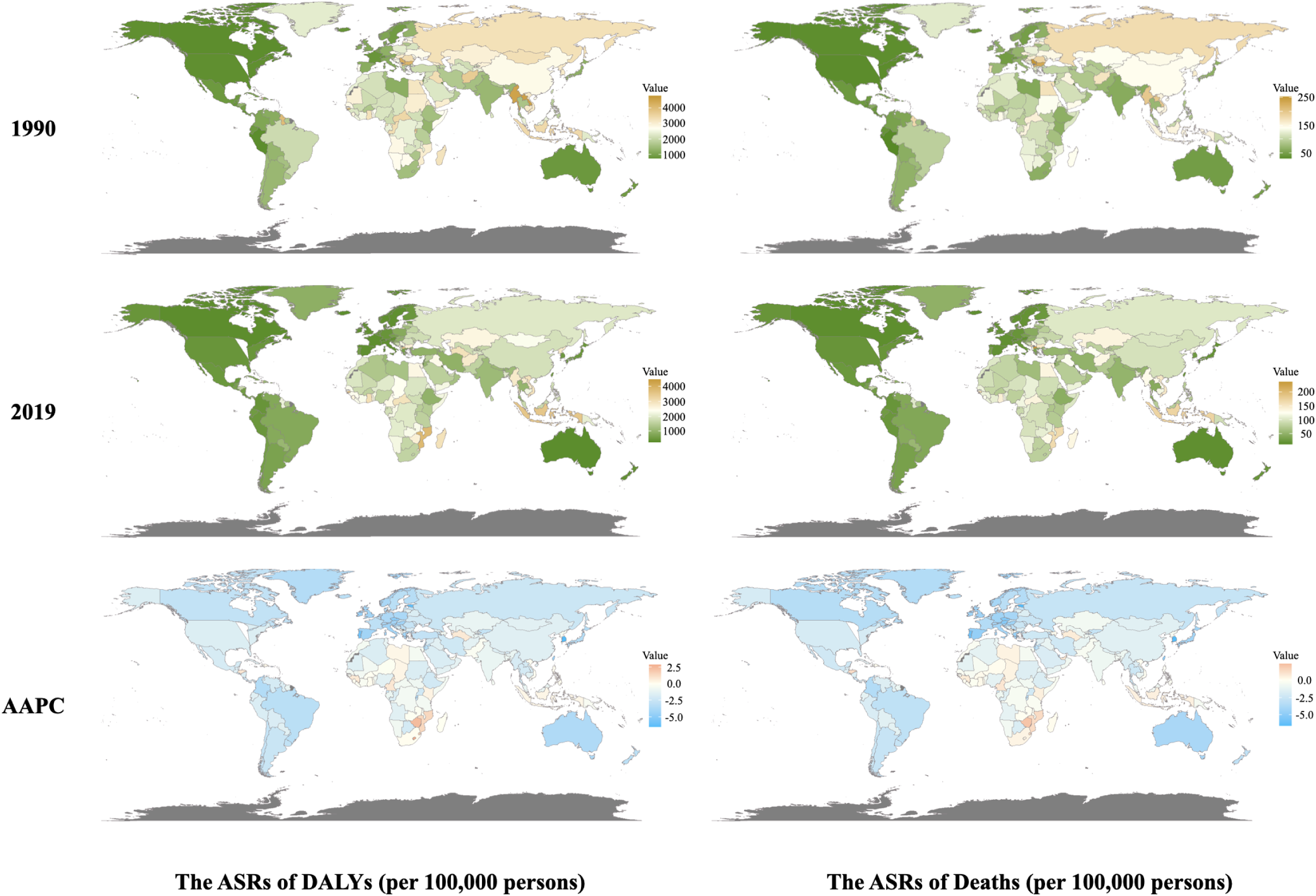
The Global Distribution of Metabolic-related Stroke from 1990 to 2019 in Country Level. ASR: Age-Standardized Rate; AAPC: Average Annual Percent Change. ASR of Deaths (per 100,000 persons) refers to the Age-Standardized Rate of deaths per 100,000 persons; ASR of DALYs (per 100,000 persons) refers to the Age-Standardized Rate of DALYs per 100,000 persons; AAPC: The Average Annual Percent Change between 1990 to 2019 in 204 countries around the world.1990: Age-standardized mortality and DALYs rates for metabolic-related stroke in 204 countries around the world in 1990; 2019: Age-standardized mortality and DALYs rates for metabolic-related stroke in 204 countries around the world in 2019.

**Figure 2:**
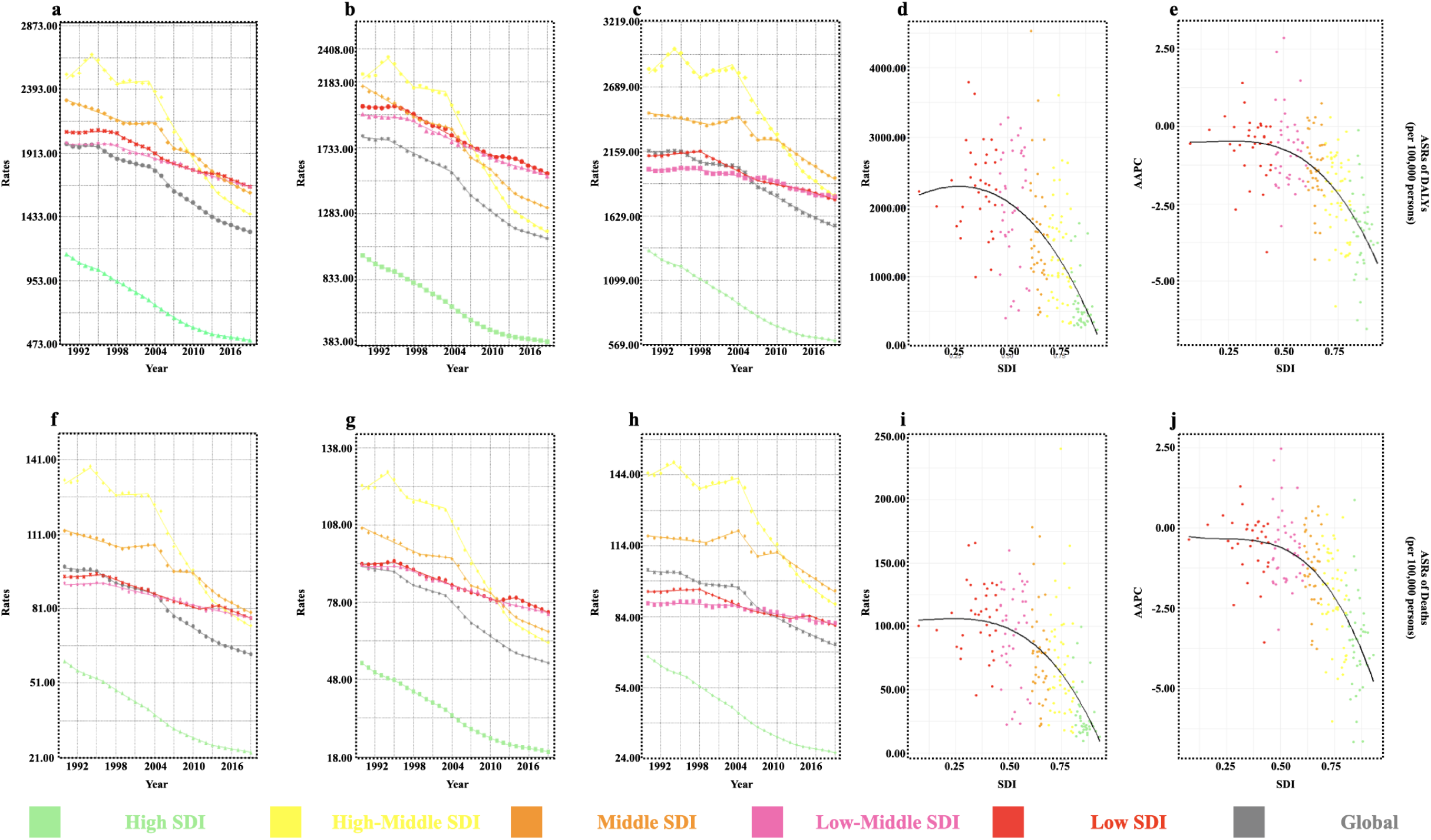
Age-Standardized Mortality Rates and DALYs for Metabolic-Related Stroke by SDI, 1990-2019. ASR: Age-Standardized Rate; AAPC: Average Annual Percent Change. ASR of Deaths (per 100,000 persons) refers to the Age-Standardized Rate of deaths per 100,000 persons; ASR of DALYs (per 100,000 persons) refers to the Age-Standardized Rate of DALYs per 100,000 persons; AAPC: The Average Annual Percent Change between 1990 to 2019. a, f: Age-standardized DALYs rate(a) and ASMR (f) for metabolic-related stroke by 5 SDI level during 1990-2019 in both genders. b, g: Age-standardized DALYs rate(b) and ASMR (g) for metabolic-related stroke by 5 SDI level during 1990-2019 in female. c, h: Age-standardized DALYs rate(c) and ASMR (h) for metabolic-related stroke by 5 SDI level during 1990-2019 in male. d, i: Age-standardized DALYs rate (d) and ASMR (i) for metabolic-related stroke by SDI around 204 countries during 1990-2019 in both genders. e, j: AAPC of Age-standardized DALYs rate(e) and ASMR (j) for metabolic-related stroke by SDI around 204 countries during 1990-2019 in both genders.

### 3.2 Impact of SDI on Stroke Burden Attributable to Metabolic Risks

We found that in 2019, regions with high SDI had significantly lower ASMR (23.15 [18.51, 26.85]) and DALYs ASR (504.31 [417.8, 582.61]) compared to other SDI regions (Table 1, Figure 2a). The AAPC was also the lowest in five SDI level regions, with AAPC of mortality at -3.47% (−3.61%, -3.32%) and DALYs at -3.03% (−3.17%, -2.89%) (Table 1). However, regardless of whether it was in 1990 or 2019, the regions with the highest ASR of DALYs or mortality rates were not the low SDI regions (Table 1). The regions with high-medium SDI exhibited a significant upward trend in both DALYs and mortality rates from 1990 to 1994 (Figure 2 a, f).

Among 204 countries, we found that as SDI increased, both ASR of DALYs (Figure 2d) and mortality rates (Figure 2i) in 2019 showed a significant decline. The AAPC from 1990 to 2019 also displayed a downward trend with the SDI upward (Figure 2e, j), and this decline was exponential; the higher the SDI, the more pronounced the decrease (Figure 2d, e, i, j).

### 3.3 Gender and Age Heterogeneity in Stroke Burden Due to Metabolic Risks

Globally, the impact of metabolic risks on males was greater than on females both on DALYs ASR and ASMR, and the trend of change was consistent for both genders, with AAPCs being less than 0 (Table 1). Except for the low-middle SDI region, where the ASMR of male was lower than that of female during 1990-1999, male consistently had higher age-standardized mortality rates and DALYs rates in all other SDI level regions and time periods during 1990-2019. (Figure 2b, c, g, h).

In different age groups, the stroke-related DALYs and deaths rates predominantly affected individuals aged 75 and above, with a lesser impact on those under 55. (Figure 3a). For the 20-24 age group, the AAPC from 1990 to 2019 was greater than 0 for both DALYs 5.18 (4.59, 5.78) and mortality rates 5.84 (5.07, 6.61) (Figure 3b), but the 2019 rates of DALYs 7.40 (0.52, 16.74) and deaths 0.08 (0.003, 0.18) was visible lower than other age groups (Figure 2a). All other age groups showed significant declines (Figure 3b). Additionally, as the SDI level increased, the percentage of DALYs or mortality rates among the population over 85 years old rose compared to other age groups. In North America, high-income Asia Pacific, Western Europe, and Australasia, individuals over 85 accounted for more than 70% of the 2019 mortality rate from metabolic-related strokes (Figure 3c).

**Figure 3:**
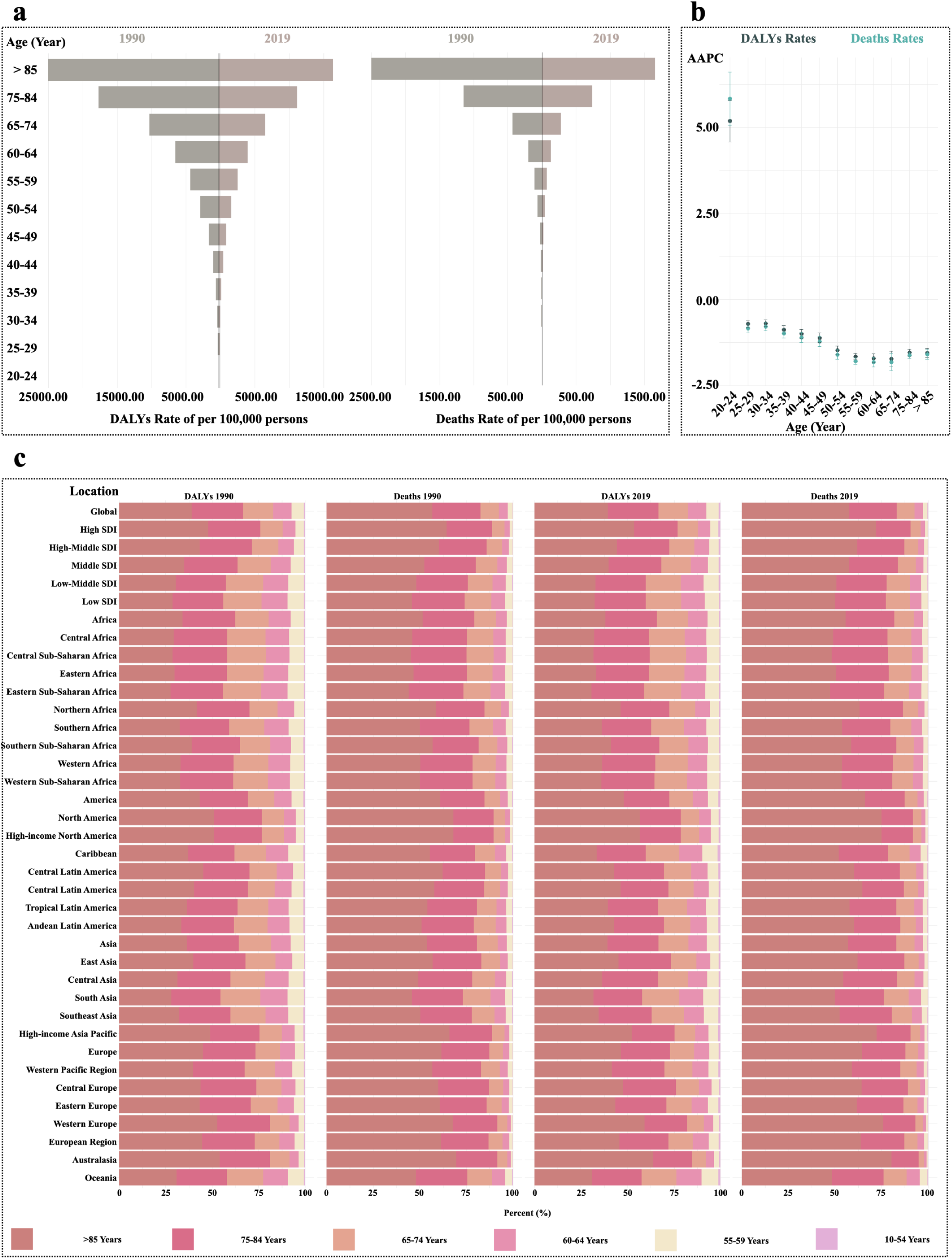
Mortality Rates and DALYs for Metabolic-Related Stroke by Age, 1990-2019. SDI: Socio-demographic Index; AAPC: Average Annual Percent Change. Death rate of per 100,000 persons refers to the Age group special rate of deaths per 100,000 persons; DALYs rate of per 100,000 persons refers to the Age group special rate of DALYs per 100,000 persons; AAPC: The Average Annual Percent Change between 1990 to 2019.

### 3.4 Proportional Contribution of Risk Factors to Stroke

Comparing age-standardized mortality and DALYs due to the three primary risk factors identified in the GBD study (Environmental/Occupational risks, Behavioral risks, and Metabolic risks), we found that metabolic risks contributed the highest percentage to stroke-related ASMR (50.82%) and DALYs (48.80%) globally in 2019 (Figure 4a), followed by Environmental/Occupational risks. The 1990 composition ratio ranking was also similar to 2019 (Figure 4a). As SDI increased, the composition ratio of DALYs ASR and ASMR due to Environmental/Occupational risks decreased, while Behavioral and Metabolic risks increased (Figure 4a). In high SDI regions, metabolic risks accounted for 58.59% of stroke-related deaths in 2019 (Figure 4a). This trend was also observed across different genders (Figure 4b, c). For males, the proportion of behavioral risks was higher than for females regardless of the data collection year or outcome measure, while the proportion of metabolic and environmental risks was lower for females (Figure 4d).

**Figure 4:**
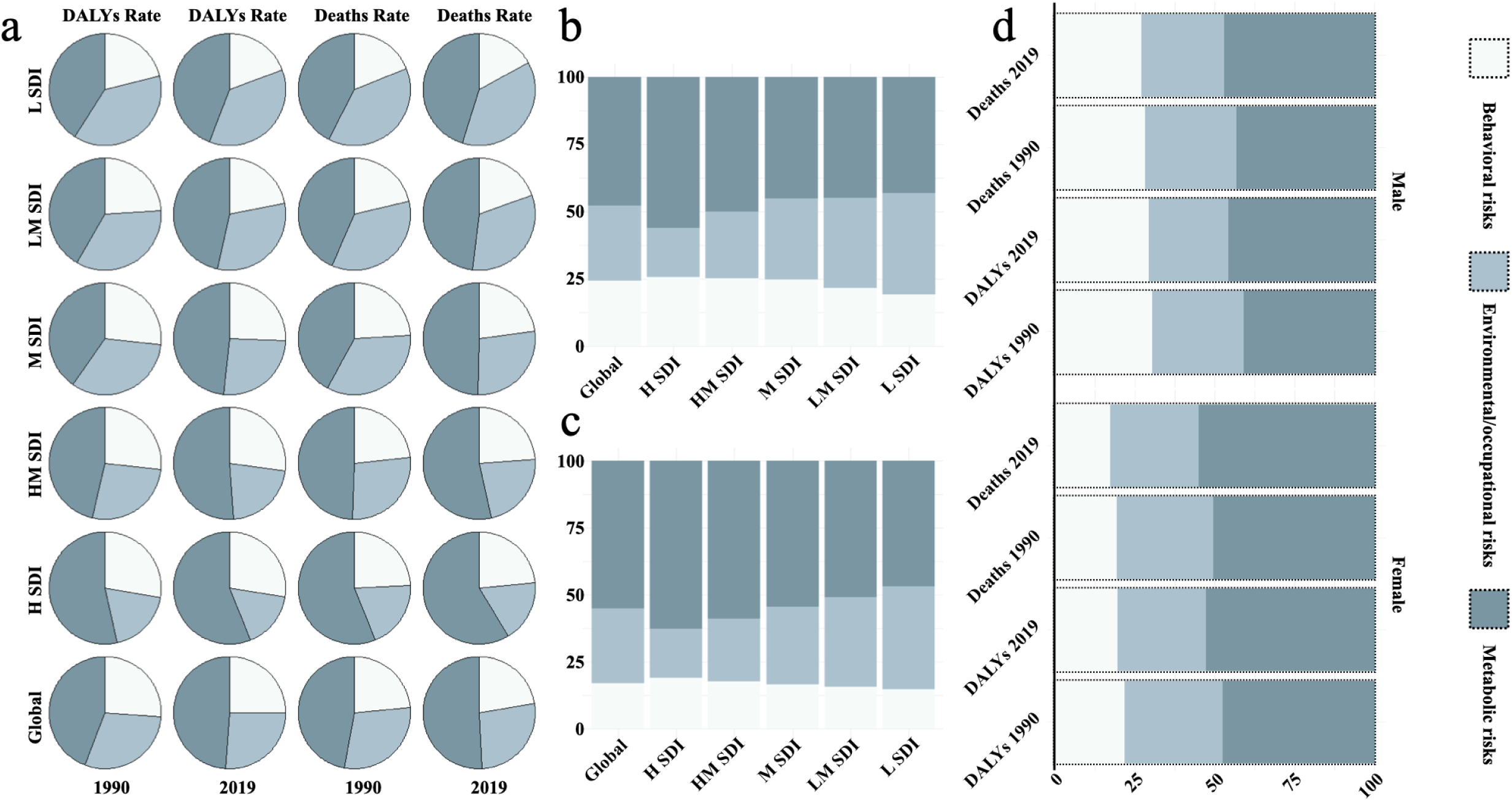
Mortality Rates and DALYs for Stroke by Risk Factors in 2019. SDI: Socio-demographic Index; H SDI: High SDI; HM SDI: High-middle SDI; M SDI: Middle SDI; LM SDI: Low-middle SDI; L SDI: Low SDI; DALYs rate: the Age-Standardized Rate of DALYs per 100,000 persons; Deaths rate: the Age-Standardized Rate of deaths per 100,000 persons. a: The proportion of contributions from different risk factors to stroke varies across years, SDI level, and outcome indicators. b, c: The proportion of contributions from different risk factors to the ASMR of stroke in 2019 varies across gender (b: male; c: female) and SDI level. d: The proportion of contributions from different risk factors to stroke varies across years, gender, and outcome indicators.

### 3.5 Burden Proportions of Stroke Types Attributable to Metabolic Risks

Based on the GBD study’s four-level causes, stroke was categorized into intracerebral hemorrhage, ischemic stroke, and subarachnoid hemorrhage. ^15^ From 1990 to 2019, age-standardized mortality and DALYs rates for these stroke subtypes attributable to metabolic factors showed a declining trend globally and across all SDI regions (Figure 5a, b). The most significant decline in DALYs for ischemic stroke was observed in high-middle SDI regions, dropping from 1478.48 (1276.71, 1648.34) in 1990 to 869.25 (727.48, 981.48) in 2019 (Figure 5a, b).

**Figure 5:**
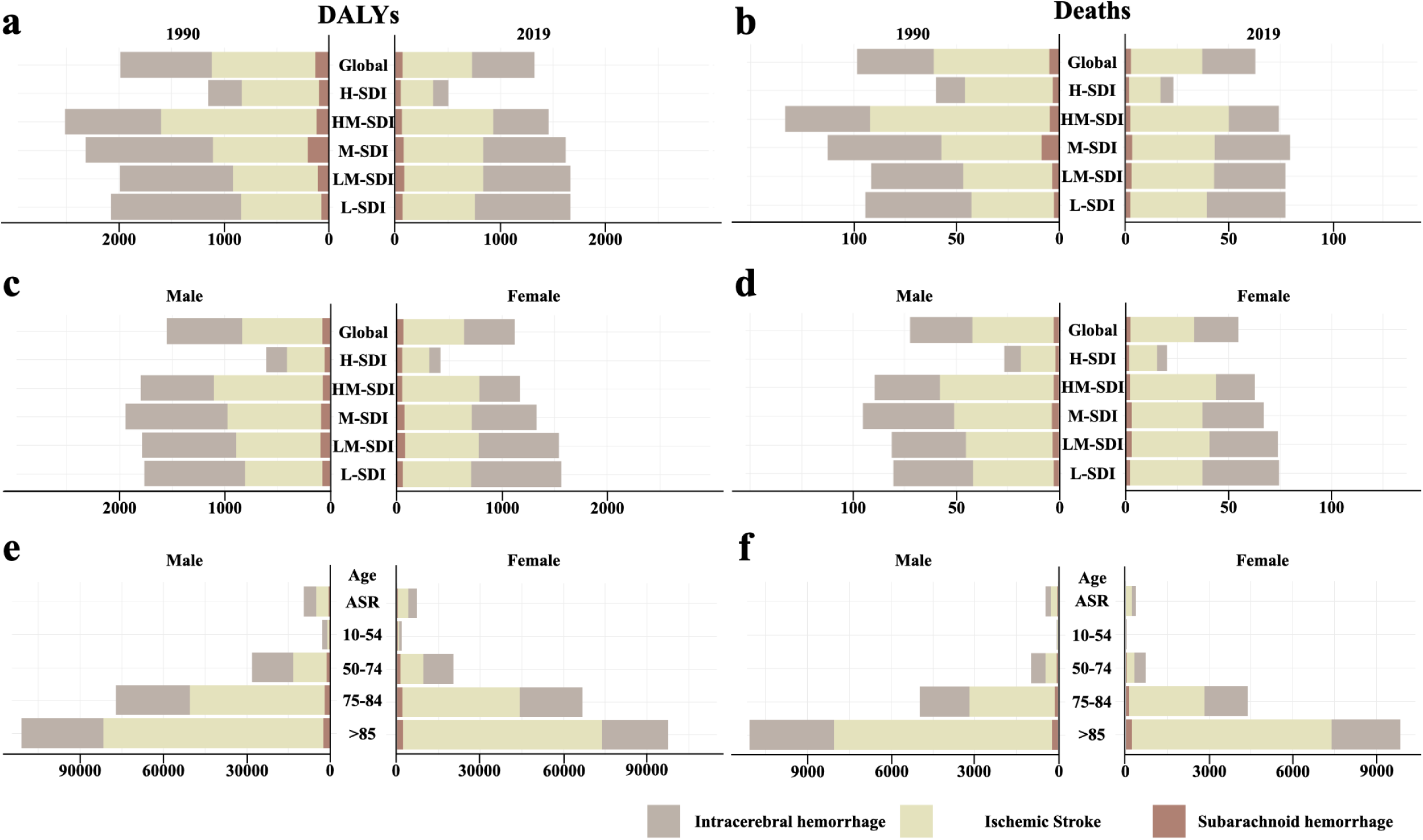
Mortality Rates and DALYs for Subgroup of Stroke in 1990 and 2019. SDI: Socio-demographic Index; H SDI: High SDI; HM SDI: High-middle SDI; M SDI: Middle SDI; LM SDI: Low-middle SDI; L SDI: Low SDI; DALYs rate: the Age-Standardized Rate of DALYs per 100,000 persons; Deaths rate: the Age-Standardized Rate of deaths per 100,000 persons. a, b: The ASR of DALYs (a) and deaths (b) of stroke different subtypes varies across years and SDI level. c, d: The ASR of DALYs (c) and deaths (d) of stroke different subtypes in 2019 varies across gender and SDI level. e, f: The age specific rate of DALYs (e) and deaths (f) of stroke different subtypes varies across gender and age group.

Globally in 2019, ischemic stroke due to metabolic risks had the highest AMSR (34.67 [28.44, 39.67]) and DALYs rate (661.26 [554.38, 749.68]) among the three subtypes, while subarachnoid hemorrhage had the lowest AMSR (2.41 [1.79, 3.03]) and DALYs rate (67.61 [49.87, 85.50]) (Figure 5a, b). With decreasing SDI levels (from High to Low), the ASMR for intracerebral hemorrhage increased (Figure 5b). In high SDI regions, the ASMR for ischemic stroke was 15.11 (12.20, 17.67) compared to 6.44 (5.20, 7.66) for intracerebral hemorrhage, and the rates were 37.20 (29.70, 45.89) for ischemic stroke and 38.02 (29.19, 46.39) for intracerebral hemorrhage in the low SDI regions (Figure 5b). Except for the slightly lower age-standardized mortality rate for subarachnoid hemorrhage in high SDI regions for males (1.55 [1.13, 1.89]) compared to females (1.61 [1.16, 2.00]), age-standardized DALYs and mortality rates were higher in males for all subtypes globally and across SDI regions (Figure 5c, d). For both genders, the 10-55 and 50-74 age groups had the highest DALYs and mortality rates due to intracerebral hemorrhage among the three subtypes (Figure 5e, f). For those aged 75-84 and over 85, ischemic stroke was the leading subtype contributing to DALYs and mortality rates (Figure 5c, d). Across all age groups, subarachnoid hemorrhage had the lowest DALYs and mortality rates among the three subtypes (Figure 5c, d).

## Discussion

Despite the similarities in global distribution patterns between metabolic risk related stroke and stroke caused by all factors ^15^—such as overall global levels and the decreasing ASMR and DALY rates across the five SDI levels—there are notable differences in how metabolic risk-related stroke is distributed by region and SDI compared to stroke from all causes. ^15^ For example, in 2019, the highest stroke mortality rate from all causes was observed in the low SDI category, while the lowest incidence of metabolic stroke occurred in the low-middle SDI category. Oceania had the highest all-cause stroke-related DALYs in 2019, with a rate of 2,998.97 (2,432.79, 3,617.12). In contrast, Southeast Asia had the highest metabolic stroke-related DALYs, with a rate of 2,500.54 (2,104.75, 2,894.53).

Southeast Asia had the highest ASMR for metabolic stroke among the 33 regions, while Singapore, also located in Southeast Asia, had the lowest ASMR for metabolic stroke in 2019. (Supplemental Table 2) This suggests that ethnicity and geographic location may not be the primary factors influencing metabolic stroke mortality or DALY rates. ^2^ ^27^ Instead, public health and healthcare systems, socioeconomic factors, and lifestyle choices likely play a more significant role. ^28^ ^29^ From 1990 to 2019, South Korea saw the most significant decline in the ASR of metabolic stroke DALYs, while Estonia experienced the most notable decrease in mortality rates. ^30^ During this period, South Korea’s Gross Domestic Product (GDP) per capita rose from $6,000 in 1990 to $31,500 in 2019 from World Bank website. Similarly, Estonia underwent significant socioeconomic improvements after gaining independence from the Soviet Union in 1991, transforming into a high-income country with an efficient economic and social system. ^30^

In contrast, sub-Saharan Africa and Southern Africa did not see significant declines in metabolic stroke rates (Table 1). This could be due to metabolic risk factors often being overlooked in African stroke patients, with the overall stroke proportion not decreasing. ^31^ ^32^ ^33^ Furthermore, Southern Africa has the highest prevalence of metabolic syndrome in Africa. ^34^ Addressing metabolic risk factors is likely the most effective way to mitigate the impact of strokes in Africa. ^29,35^ Lesotho, another Southern African country, saw the most significant increase in both ASR of DALYs and deaths from 1990 to 2019 (Supplemental Table 2). Clinical studies indicate that CVDRF is highly prevalent across all age and gender groups in Lesotho, ^36^ highlighting the importance of strengthening preventive measures in Lesotho and similar regions in Southern Africa. Overall, enhancing national economic levels, ^37^ developing plans to protect against and manage metabolic risk factors, ^38^ and individuals paying more attention to metabolic factors related to stroke^38^ were crucial for reducing DALYs and mortality rates caused by metabolic stroke.

As SDI increases, ASR of DALYs and mortality rates shown a similar exponential decline, and the AAPC trend is consistent. This indicates that developing countries with weak socioeconomic foundations face a greater challenge of a sustained burden of metabolic stroke compared to developed nations. ^39^ In high-income regions such as North America, high-income Asia Pacific, Western Europe, and Australia, individuals aged 85 and above accounted for over 70% of metabolic stroke mortality in 2019. These high-income countries face more severe aging problems, ^40^ with the proportion of elderly people increasing annually. ^40^ As people age, they accumulate metabolic-related diseases (such as diabetes, hypertension, and high cholesterol) ^41^, increasing their risk of stroke. Additionally, the physiological frailty of individuals aged 85 and above results in poorer stroke outcomes. ^42^ These findings suggest the need to strengthen the prevention and management of metabolic-related diseases in developing countries. In high-income countries, should pay more attention to the issue of population aging.

Our study also found that, regardless of data collection year and outcome measures, the proportion of behavior risk-related stroke is consistently higher in men than in women, while the proportion of metabolic and environmental risk-related stroke is lower in men. However, strokes related to these three risk factors result in more severe DALYs and mortality rates in men. This may be due to men’s insufficient management of metabolic-related diseases (such as hypertension and diabetes), combined with the additive effect of high behavioral risks, ^43^ leading to a higher risk of stroke and more severe outcomes. Increasing men’s awareness and management of metabolic diseases, along with regular health screenings and early intervention, can lower stroke incidence and severity. ^44^

## Limitations

Despite the comprehensive nature of our analysis, several limitations should be acknowledged. The reliance on secondary data sources may introduce reporting biases. The variability in healthcare infrastructure and diagnostic capabilities across regions could affect the accuracy of our findings.

## Conclusion

Our study highlights significant global progress in reducing the stroke burden attributable to metabolic risks, particularly in high SDI regions. However, persistent disparities across different SDI levels, genders, and age groups necessitate continued efforts to address these gaps. Comprehensive public health strategies targeting metabolic risks, with a focus on high-risk populations, are essential to further mitigate the global stroke burden.

## Disclosure

The authors declare no conflict of interest.

## Data Availability

Data used in the analyses can be obtained from the Global Health Data Exchange Global Burden of Disease Results Tool (https://ghdx.healthdata.org/gbd-results-tool

